# Predicting 7-Day Re-Bleeding after Peptic Ulcer Hemostasis: Retrospective Analysis of Forrest, Complete Rockall, and Glasgow-Blatchford Scores and Independent Risk Factors

**DOI:** 10.64898/2025.12.10.25342008

**Authors:** Niranta Kumar Das, Md Abeed Hasan, Sayed Abdullah Jami, Charls Erik Halder

## Abstract

**Objective:** To identify independent risk factors for early (≤7-day) re-bleeding after peptic ulcer bleeding (PUB) and to compare the predictive performance of Forrest classification, Complete Rockall Score (CRS), and Glasgow-Blatchford Score (GBS).

**Methods:** We retrospectively analyzed adults with endoscopy-confirmed peptic ulcer bleeding from 2015–2020. Early re-bleeding was defined as ≤7 days after index hemostasis. We applied univariable and multivariable logistic regression and assessed discrimination with ROC curves (AUC).

**Results:** Independent risk factors for early re-bleeding included: heart rate (OR 1.054), hemoglobin (OR 1.878), erythrocyte distribution width (OR 1.171), degree of ulcer erosion (OR 1.191), and blood transfusion intervention (OR 12.296). Forrest showed the best discrimination (AUC 0.775; sensitivity 96.2%; specificity 58.8%), followed by GBS (AUC 0.670) and CRS (AUC 0.507)

**Conclusions:** Heart rate, hemoglobin, erythrocyte distribution width, ulcer erosion, and blood transfusion are significant risk factors for early re-bleeding in PUB. Forrest grading is the most effective predictor, while GBS can stratify risk and may benefit from modifications. CRS showed limited predictive utility.

**Limitations:** single-center, retrospective design; possible residual confounding; no external validation.

**Clinical implications:** Forrest can guide intensified monitoring/hemostasis; GBS supports pre-endoscopy triage; CRS adds limited value.

## INTRODUCTION

Peptic ulcer and bleeding peptic ulcer (PUB) are common and potentially life-threatening conditions encountered in emergency and gastroenterology departments. The primary clinical manifestations include hematemesis (vomiting blood) and melena (black or tarry stools). Approximately 20%-30% of patients with ulcers experience bleeding. Severe cases, involving bleeding volumes of 1000 ml or a reduction of blood volume exceeding 20%, can rapidly lead to hemorrhagic shock and significant mortality, accompanied by a high rate of re-bleeding [1]. Notably, the incidence rate of PUB varies globally, with a particularly high prevalence in China [2]. Several studies [3] report that the re-bleeding rate after PUB treatment in China ranges from 14%–36%, with a mortality of 6%–10%, higher than in Europe and North America. Patients with adverse clinical factors face a significantly increased risk of early re-bleeding, leading to greater medical burdens and diminished quality of life. Several studies [4–11] have identified risk factors for early re-bleeding in PUB, including patient demographics (e.g., age, sex), clinical indicators (e.g., haemoglobin, serum albumin, blood urea nitrogen), and ulcer features (e.g., size, depth, Helicobacter pylori). These risk factors, however, remain the subject of ongoing debate across different clinical studies. This study expands on prior research by analyzing a larger sample size and incorporating multiple risk factors. Its primary objectives are to identify independent predictors of early re-bleeding in PUB and to guide individualized treatment strategies aimed at reducing re-bleeding risk and improving patient outcomes. To predict early clinical outcomes in PUB patients, effective and reliable scoring systems are critical. Internationally recognized systems include the Rockall re-bleeding risk score, Glasgow-Blatchford Score (GBS), AIMS65, the Baylor bleeding score, and the APACHE scoring system [12]. Among these, the Forrest classification remains widely recommended for its endoscopic description of lesions and utility in predicting re-bleeding risk [13–14]. The Rockall score integrates clinical symptoms, laboratory data, and endoscopic findings, offering a comprehensive approach for predicting re-bleeding and mortality [15]. Similarly, the GBS, based on clinical symptoms and laboratory data, is particularly valuable for stratifying re-bleeding risk before endoscopic evaluation and predicting surgical interventions. Despite their utility, these scoring systems have unique strengths and limitations, which have led to some clinical controversies regarding their prognostic value. Recent studies indicate that endoscopic signs such as congestion, edema, and erosion around ulcers—often triggered by non-steroidal anti-inflammatory drugs or stress—are significant risk factors for re-bleeding in PUB patients. Many patients with severe PUB require urgent treatment due to massive bleeding from erosion and active oozing of blood. This study analyzes the clinical, laboratory, and endoscopic data of PUB patients, utilizing the Forrest grading system, Rockall score, and GBS to evaluate their predictive value for early re-bleeding. The findings aim to provide valuable insights for improving clinical diagnosis and management of PUB.

## MATERIALS AND METHODS

### Study Design, Participants, Inclusion and Exclusion Criteria

This study analyzed patients treated for peptic ulcer bleeding (PUB) at the General Hospital of Ningxia Medical University from May 2015 to May 2020. All diagnoses were confirmed by endoscopy, and patients received omeprazole acid suppression therapy after admission. Inclusion criteria required patients to be aged ≥18, exhibit symptoms of gastrointestinal bleeding (e.g., hematemesis or melena), and have a gastroscopic diagnosis of PUB, following the *Guidelines for Acute Non-variceal Upper Gastrointestinal Bleeding (ANVUGIB)* [14].Exclusion criteria included inability to undergo endoscopy, incomplete data for Forrest grading, Rockall Score (RS), or Glasgow-Blatchford Score (GBS), discontinuation or transfer during treatment, bleeding from non-PUB causes, or conditions such as malignancy, pregnancy, or severe multi-organ disease. These ensured a focused and reliable study population.

### Inclusion Criteria

Adults with endoscopy-confirmed PUB; index admission; complete outcome data for 7-day re-bleeding.

### Exclusion Criteria

Variceal bleeding; malignant ulcers; incomplete key covariates; lost to 7-day follow-up.

### Diagnostic Criteria

The diagnosis of peptic ulcer bleeding (PUB) is established according to the *Guidelines for Diagnosis and Treatment of Acute Non-variceal Upper Gastrointestinal Bleeding (ANVUGIB)* when two primary criteria are met. First, patients must exhibit symptoms such as hematemesis, melena, dizziness, pale complexion, increased heart rate, and decreased blood pressure, indicative of significant gastrointestinal bleeding. Second, endoscopic examination must confirm the absence of esophageal or gastric fundus varices while identifying a bleeding lesion within the upper gastrointestinal tract. Early re-bleeding in PUB is diagnosed if specific clinical and laboratory criteria are observed within one week after the cessation of bleeding [14]. Clinically, increased episodes of hematemesis or melena, characterized by bright red vomitus or red to dark red fecal discharge, are warning signs. Laboratory findings, such as a hemoglobin drop of more than 20 g/L within 24 hours or hemoglobin levels persistently below 10 g/L despite multiple transfusions, indicate ongoing bleeding. Signs of peripheral circulatory failure, including systolic blood pressure below 90 mmHg or a pulse rate exceeding 110 beats per minute, also signify re-bleeding. If circulatory status worsens despite adequate fluid replacement and blood transfusion or initially improves but then deteriorates (e.g., decreasing hemoglobin, red blood cell count, and hematocrit alongside an increasing reticulocyte count), re-bleeding should be suspected. Additionally, despite adequate fluid and urine output, persistent or rising blood urea nitrogen levels suggest recurrent bleeding. Finally, re-bleeding is confirmed when repeat gastroscopy identifies active bleeding at the base of the ulcer. These criteria provide a structured approach to identifying early re-bleeding, enabling timely clinical intervention.

### Groups

According to the patient’s medical history, physical signs, auxiliary examinations and gastroscopy, patients with re-bleeding are assessed and grouped. After conservative medical treatment and endoscopic treatment, the patient’s vital signs were stable; symptoms improved and no bleeding manifested.

### Scoring System and Shock

See in Figure 1, Table 1,2,3. Evaluation criteria for the severity of the erosion around the ulcer according to the 2004 "Trial Opinions on Endoscopic Classification and Grading Standards and Treatment of Chronic Gastritis” such as A is indicated in red, B is indicated black and C represents surface cleanliness.

**Table 1.** FORREST CLASSIFICATTON.

| **FORREST GRADING** | **ULCER LESIONS** | **FORREST GRADING** | **ULCER LESIONS** |
| --- | --- | --- | --- |
| IA | Active hemorrhage | IIB | Adherent clot |
| IB | Oozing hemorrhage | IIC | Flat pigmented clot (Black base) |
| IIA | Non bleeding visible vessel | III | Clean-based ulcer & no signs of recent bleeding |

**Table 2.** COMPLETE ROCKALL (CRS) SCORING SYSTEM.

| **Variable** | **Score- 0** | **Score 1** | **Score 2** | **Score 3** |
| --- | --- | --- | --- | --- |
| Age | <60 | 60-79 | ≥80 | ----- |
| Shock status | No shock | Tachycardia | Hypotension | ----- |
| Co-morbidity | Nil Major | ----- | Congestive cardiac failure , Ischemic Heart Disease, Major morbidity | Renal or liver metastatic cancer |
| Endoscopic Diagnosis | Mallory Weise tear | All other diagnoses | Malignancy of the upper GI tract | ----- |
| Evidence of endoscopic Bleeding | None | ----- | Blood adherent clot, spurting vessel. | ----- |

**Table 3.** GLASGOW BLATCHFORD SCORING (GBS) SYSTEM.

| **CLINICAL PARAMETER** | **Range** | **Score** |
| --- | --- | --- |
| Hemoglobin for men (g/l) | 120-----<129 | 1 |
|  | 100-----<119 | 3 |
|  | <100 | 6 |
| Hemoglobin for women (g/l) | 100-----<1119 | 1 |
|  | <100 | 6 |
| Blood urea nitrogen (mmol/l) | 6.5---<7.9 | 2 |
|  | 8.0----<9.9 | 3 |
|  | 10.00----<24.9 | 4 |
|  | ≥25.0 | 6 |
| Systolic blood pressure(mmHg) | 100-109 | 1 |
|  | 90---99 | 2 |
|  | <90 | 3 |
| **Other markers** |  |  |
| Pulse | ≥100 | 1 |
| Presentation with melena |  | 1 |
| Presentation with syncope |  | 2 |
| Heart disease |  | 2 |
| Cardiac Diseases |  | 2 |

**Score interpretation** ≥6 are classified as medium to high risk, <6 as low risk.

### Statistical Analysis

In this study, statistical analysis was performed using SPSS version 22.0. Measurement data were expressed as the mean ± standard deviation, and comparisons between groups were conducted using an independent sample t-test. Enumeration data were represented as composition ratios or rates, and comparisons between groups were carried out using the chi-square test (χ² test). Univariate and multivariate logistic regression analyses were performed to identify related factors, with the odds ratio (OR) and 95% confidence interval (95% CI) calculated for each risk factor. The area under the curve (AUC) and 95% CI of the receiver operating characteristic (ROC) curve for each scoring system were analyzed and compared to evaluate their ability to predict re-bleeding and the prognosis of patients with peptic ulcer bleeding (PUB). A probability value of *p*<0.05 was considered statistically significant.

### Missing Data and Bias Control

Analyses used complete-case data; per-variable missingness is reported in Table S1. Sensitivity analyses excluding variables with >10% missingness yielded similar estimates.

### Confounding

Multivariable models included a priori covariates (age, sex, heart rate, haemoglobin, comorbidities, NSAID/antiplatelet use, and endoscopic stigmata) to reduce confounding.

### Inter-observer Agreement

Two experienced endoscopists assigned Forrest grades; inter-observer agreement was assessed (κ reported in Table S2).

## RESULTS

### Comparison of the general characteristics of the two groups of PUB patient

Patients were divided into the re-bleeding group (n=26, 4.0%) and the no-re-bleeding group (n=616, 96.0%). There were 642 PUB patients, who met the inclusion criteria including 539 males and 103 females with an average age of (51.46±17.0) years. There were statistically significant differences between the two groups of PUB patients in terms of history of peptic ulcer and heart rate on admission (*p<*0.05). However, there were no statistically significant differences in gender, age, history of NSAIDs, complications, value of systolic blood pressure in hospital admission, and length of stay (*p*> 0.05) as shown in table 4.

**TABLE 4:** COMPARISON OF GENERAL CONDITIONS BETWEEN THE TWO GROUPS OF PUB PATIENTS.

| Group | No. of cases | Gender (m/f) | Age (years of old) | History of peptic ulcer (Y/N) | Previous NSAIDs history (with or without) | Complications (Yes or No) | Admission systolic blood pressure(mmHg) | Admission heart rate (b/min) | Length of hospital stay (day) |
| --- | --- | --- | --- | --- | --- | --- | --- | --- | --- |
| No re-bleeding group | 616 | 518/98 | 51±17.03 | 112/504 | 118/498 | 325/29 | 125.37±19.58 | 82.51±14.32 | 6.78±3.32 |
| Early re-bleeding group | 26 | 21/5 | 52.69±16.53 | 22/4 | 6/20 | 16/10 | 122.57±27.52 | 90.96±16.11 | 7.61±3.35 |
| t/P value | - | 0.858 | 0.702 | .000 | 0.620 | 0.380 | 0.484 | 0.004 | 0.226 |
| Chi-square value | - | 0.032 | 0.387 | 16.665 | 0.246 | 0.772 | -0.700 | 2.930 | 1.238 |

### Comparison of results of auxiliary examination and gastroscopy between two groups of PUB patients

There were statistically significant differences between the two groups of PUB patients in terms of HGB, erythrocyte distribution width (RDW), Serum albumin, and degree of ulcer erosion (*p<*0.05), Nevertheless, there were no statistically significant differences in international standardized ratio, urea nitrogen and ulcer location (*p*> 0.05), as shown in table 5.

**TABLE 5:** COMPARISON OF LABORATORY DATA AND GASTROSCOPY RESULTS BETWEEN THE TWO GROUPS OF PUB PATIENTS.

| Group | Number of cases | Hemoglobin (g/dl) | Erythrocyte distribution width (%) | Serum albumin(g/l) | International normalized ratio | Urea nitrogen (mmol/l) | Ulcer site (stomach /duodenum) | Degree of erosion of ulcer surface<br>A=red surface.<br>B= black surface<br>C= clean surface |
| --- | --- | --- | --- | --- | --- | --- | --- | --- |
| No -re bleeding | 616 | 93.04±21.51 | 14.45±2.08 | 32.92±5.89 | 1.08±.042 | 7.70±4.98 | 232/366 | (178/263/175) |
| Early re-bleeding | 26 | 55.88±15.41 | 18.33±4.32 | 27.21±6.93 | 1.10±0.14 | 7.75±6.06 | 10/16 | (21/4/1) |
| Chi-square value/t value | - | -8.712 | 8.723 | -4.132 | 0.628 | 0.048 | 0.001 | 31.631 |
| t-value<br>p -value | - | 0.000 | 0.000 | 0.000 | 0.533 | 0.962 | 0.973 | 0.000 |

### Comparison of transfusion treatment and scoring system between two groups of PUB patients

There were statistically significant differences in transfusion intervention, Forrest grading evaluation and GBS evaluation scores between the two groups (*p*<0.05). There was no statistical significance in the value of CRS between the two groups of PUB patients (*p*> 0.05), as shown in Table 6. The risk of re-bleeding is higher in Forrest IB 20% compared to other grades (IA 16.66%, IIA8.57%, IIB3.31%, IIC1.92%) is tending to re-bleeding again (*p*-trend<0.001) in Forrest grading evaluation.

**TABLE 6.1:**
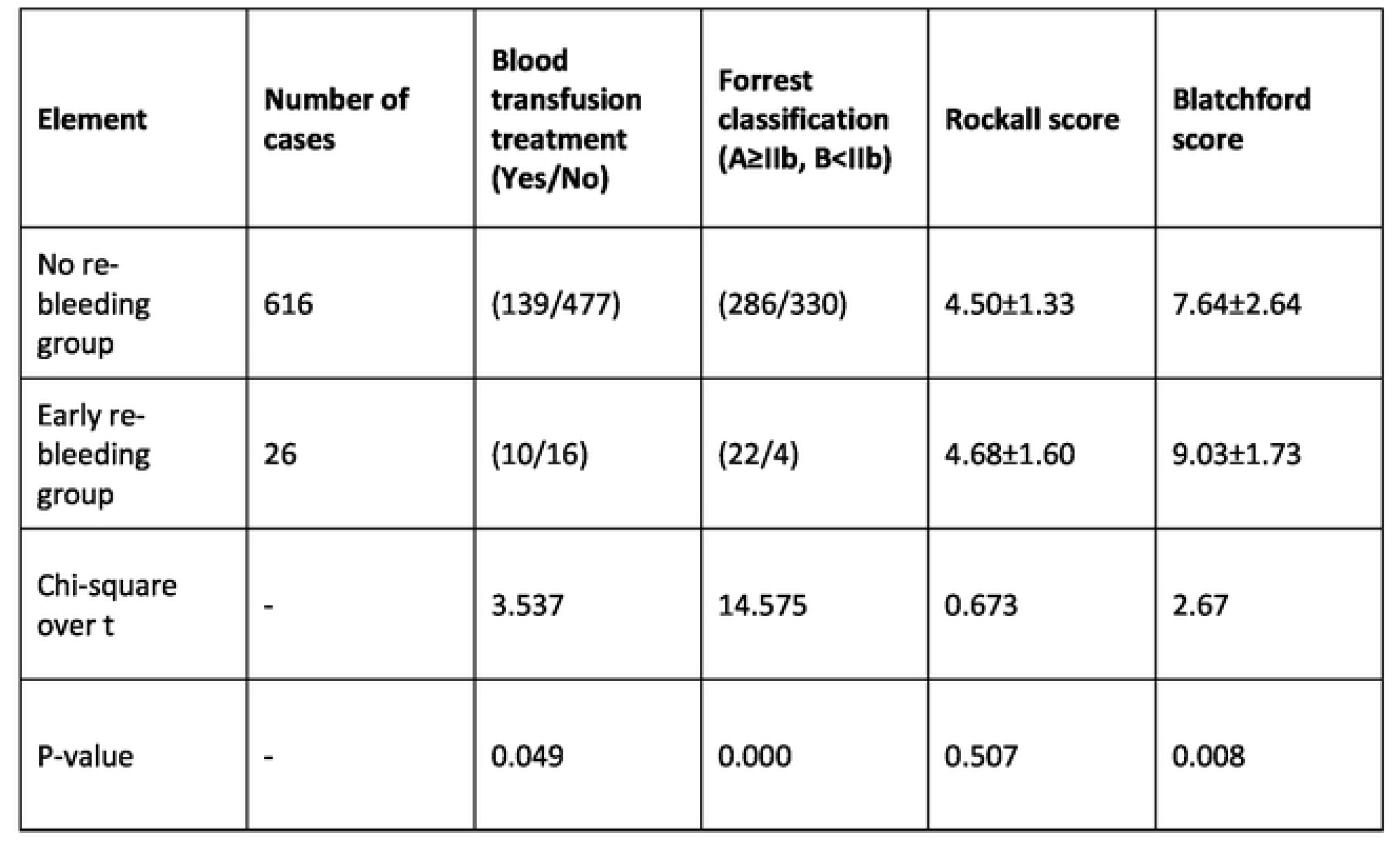
COMPARISON OF BLOOD TRANSFUSION TREATMENT AND SCORING SYSTEM BETWEEN THE TWO GROUPS OF PUB PATIENTS.

**Table 6.2:**
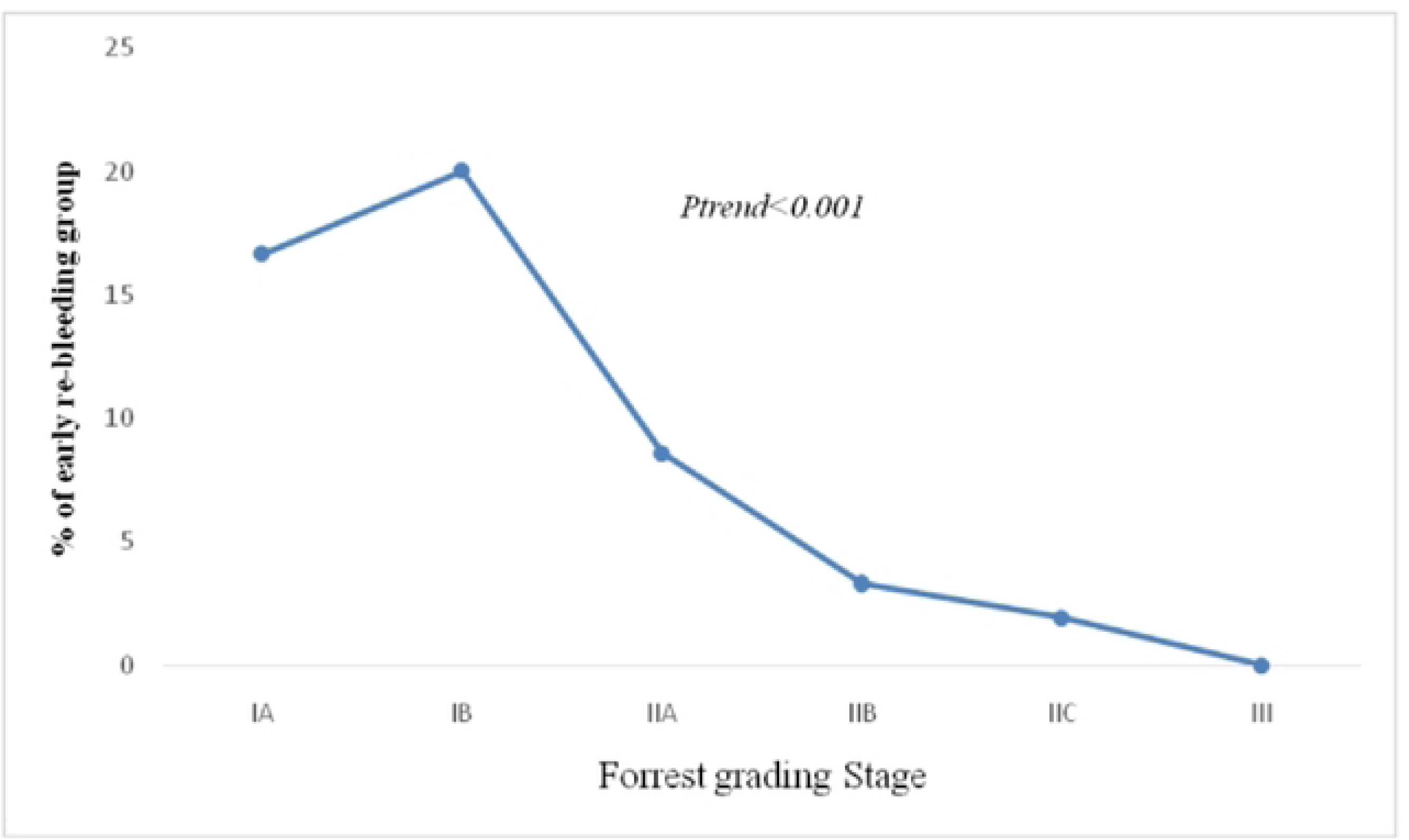
Forrest grading evaluation.

### In two groups of PUB patients, univariate Logistic regression analysis

The heart rate (OR 1.083; 95% CI 1.016–1.154), hemoglobin (OR 1.858; 95% CI 1.800–1.921), erythrocyte distribution width (OR 1.461; 95% CI 1.109–1.921), degree of ulcer-surface erosion (A/B) (OR 1.048; 95% CI 1.007–1.337), and blood transfusion intervention (OR 61.977; 95% CI 3.645–1053.813) were risk factors for early re-bleeding in PUB patients (p<0.05; Table 7).

**TABLE 7:**
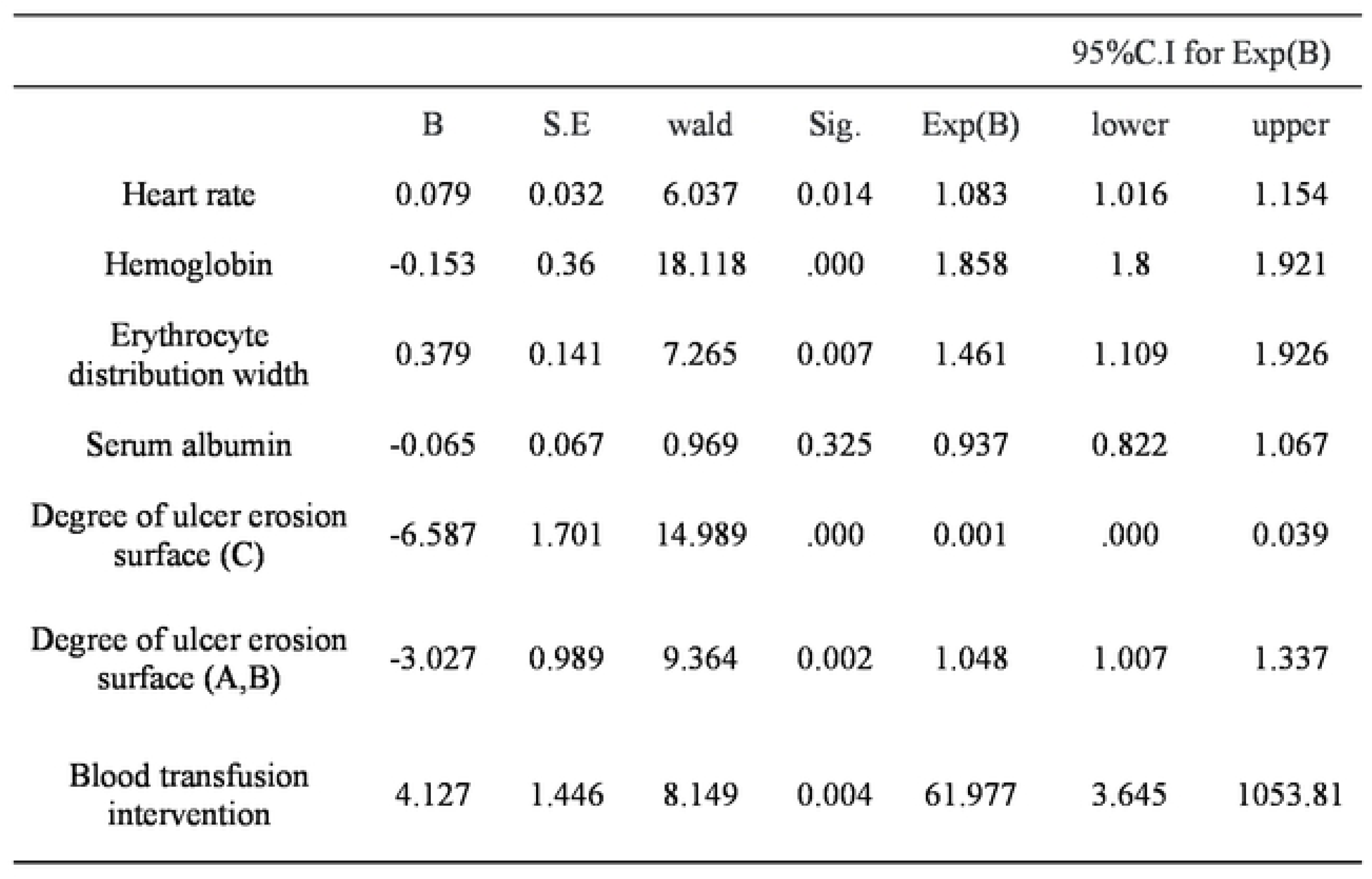
UNIVARIATE LOGISTIC REGRESSION ANALYSIS OF EARLY RE-BLEEDING IN TWO PUB GROUPS.

### Multivariate logistic regression analysis of early re-bleeding in two groups of PUB patients

Multivariable non-conditional logistic regression showed that heart rate (OR 1.054; 95% CI 1.014–1.095), hemoglobin (OR 1.878; 95% CI 1.830–1.928), erythrocyte distribution width (OR 1.171; 95% CI 0.977–1.402), ulcer-surface erosion A/B (OR 1.191; 95% CI 1.051–1.766), and blood transfusion (OR 12.296; 95% CI 1.892–79.917) were independent risk factors for early re-bleeding (Table 8).

**TABLE 8:**
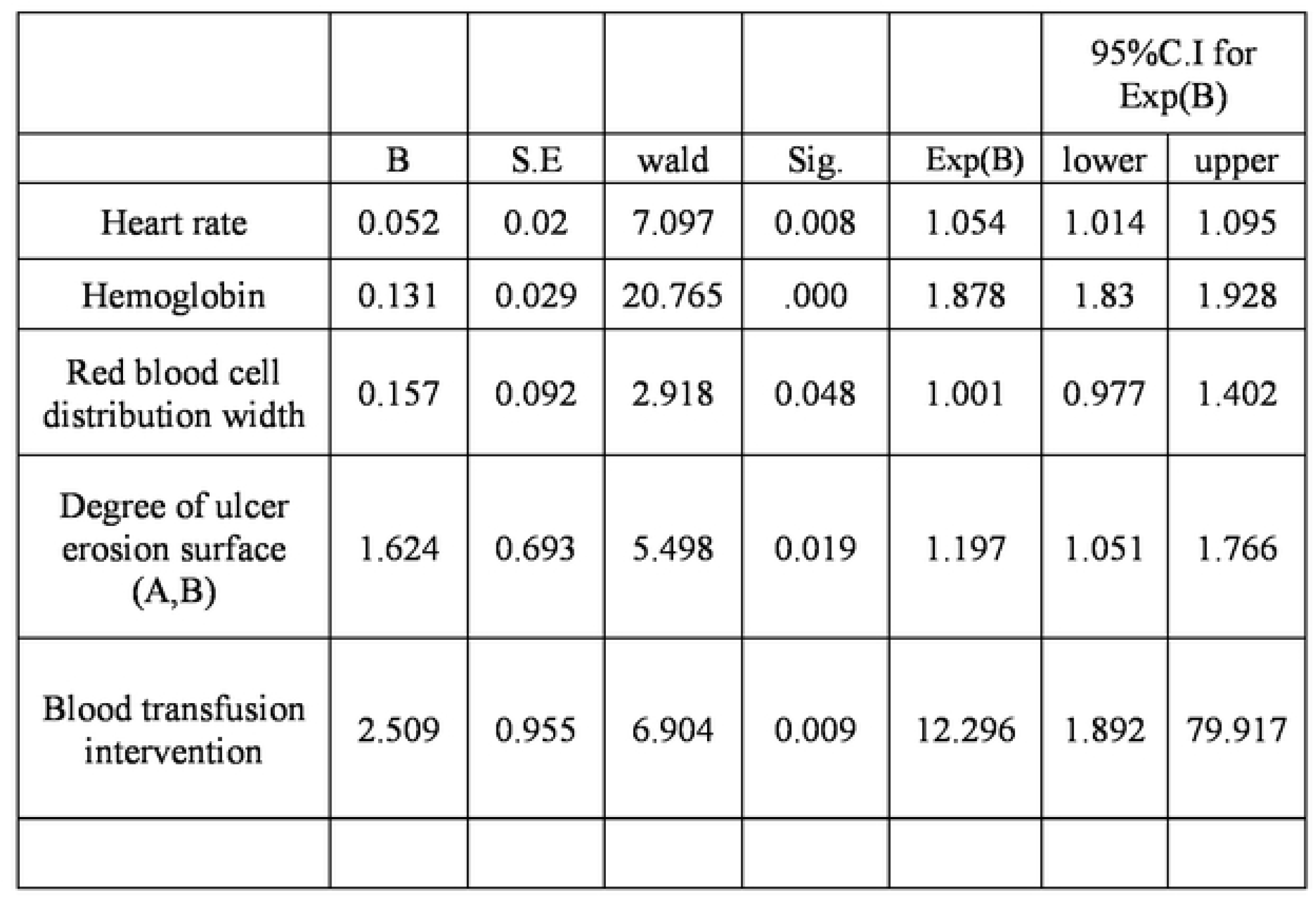
MULTIVARIATE LOGISTIC REGRESSION ANALYSIS OF EARLY RE-BLEEDI_NG IN TWO GROUPS OF PUB PATIENTS.

### Prediction of early re-bleeding in PUB patients by three scoring system

The results of predicting early re-bleeding in PUB patients with Forrest grading, GBS scoring system and CRS scoring system show that, when the Forrest classification predicts the early re-bleeding outcome of PUB patients, the area under the ROC curve is 0.775, and its sensitivity and specificity were 96.2% and 58.8% respectively. When GBS predicted PUB early re-bleeding in patients, the area under the curve was 0.670 and the sensitivity and specificity were 84.6% and 51.5%, respectively. When CRS predicted PUB early re-bleeding patients, the area under the curve was 0.507, and the sensitivity and specificity were 80.8% and 30.1%, respectively, as shown in Figure 3A–C and Tables 9–10.

**TABLE 9:** PREDICTION OF EARLY RE-BLEEDING IN PUB PATIENTS BY THREE SCORING SYSTEMS.

|  |  |  |  |  |  | Asymptotic 95%<br>confidence interval |  |
| --- | --- | --- | --- | --- | --- | --- | --- |
|  | Area | Sta.errora | Asymptotic<br>sig.b | specificity | sensitivity | Lower | upper |
| GBS<br>grading | .670 | .039 | .003 | 0.515 | 0.846 | .593 | .746 |
| Forrest<br>grading | .775 | .033 | .000 | 0.588 | 0.962 | .710 | .839 |
| CRS<br>grading | .507 | .050 | .047 | 0.301 | 0.808 | .410 | .604 |

**Table 10:**
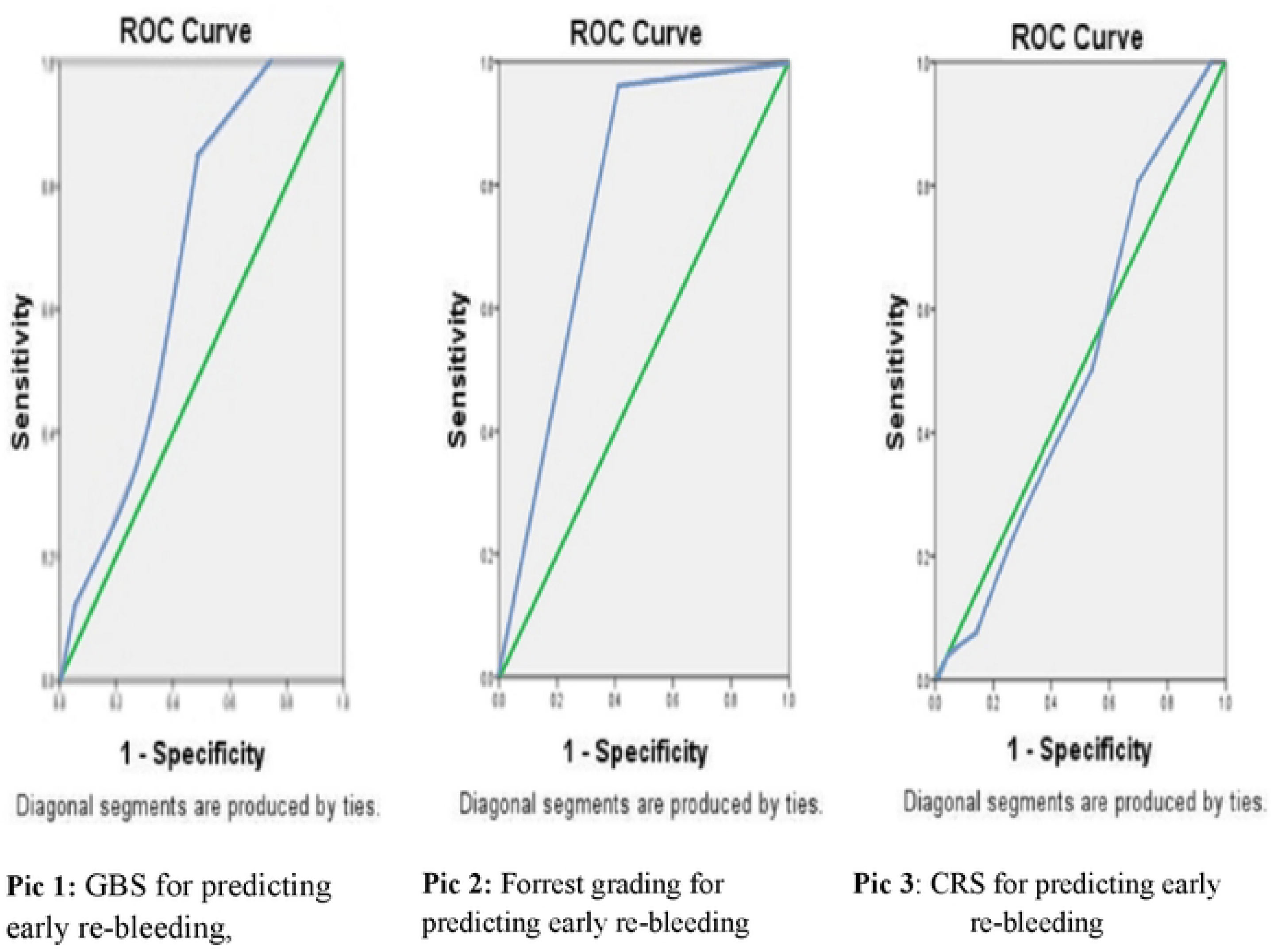
Receiver operating characteristic (ROC) curve as below.

## DISCUSSION

### Main findings

Peptic ulcer (PU) is an ulcer formed by the digestion of the lining of the gastrointestinal tract. It is often related to gastric juice acidification and digestion. Peptic ulcer (PU) is the most common cause of upper gastrointestinal bleeding, accounting for about 50%-70% of non-variceal rupture and bleeding causes in China. When peptic ulcer erodes peripheral or deep blood vessels, it may cause bleeding of different degrees. With the improvement of patient medical awareness, the wide application of proton pump inhibitors (PPI) and the continuous development of endoscopic therapeutic technique, most of patients with peptic ulcer bleeding can be cured clinically. However, the incidence of peptic ulcer bleeding (PUB) patients with re-bleeding within one week after treatment is still relatively high. Prior studies report 6.7%–22.5% short-term re-bleeding. [5–6,16] In our cohort, the rate was 4.0%, which may reflect the study population, early PPI use, and standardized endoscopic care.

### Comparison with prior studies

The etiology and pathogenic factors of patients with hemorrhage quantity differ greatly in different individual partition cells caused by excessive gastric acid output and high acid environment increased pepsinogen activation, h. pylori (Helicobacter pylori, h. pylori or Hp) infection, and long-term non-steroidal drugs (non-steroid anti-inflammatory drugs, drugs such as NSAIDs, and glucocorticoids. Mucosal defense and genetic susceptibility to repair abnormalities, such as single or interaction, can trigger various degrees of bleeding in peptic ulcer patients. In mild cases, intermittent black stools (melena) or positive fecal occulting blood may be present. In severe cases, hematemesis, dark red melena or impaired peripheral circulation may be present. Risk assessment of PUB patients is of great significance for clinical treatment and improvement of prognosis. The incidence of re-bleeding varies with different risk factors. At present, there are different reports on the risk factors of early PUB re-bleeding at domestic and abroad, which may be related to the difference of study population and clinical case data. Research reports 5 years ago showed that advanced age (> 60 years old), shock, numerous complications, nosocomial hemorrhage, low hemoglobin level, lack of follow-up PPI treatment after hemorrhage, and malignant tumor hemorrhage were associated with poor prognosis [17–19]. Peng Qionghui et al. showed that shock and concomitant liver disease were independent risk factors for predicting re-bleeding in patients [7]. Chen Chao found that patients in the observation group were older than ≥75 years old, drinking alcohol, patients with diabetes, HP infection, and previous oral NSAIDs with bleeding incidence increased compared with the control group [4]. A different study found that ulcer size and endoscopic Forrest grading were major risk factors for re-bleeding in patients [1]. There is also literature showing that besides the above factors, nutritional status, living habits, season, maintenance treatment, etc are also important factors that lead to recurrent peptic ulcer bleeding [5]. In this study, heart rate, HGB, RDW, degree of erosion of ulcer surface (A and B), and transfusion intervention were risk factors for early re-bleeding in PUB patients. Multivariate unconditional logistic regression analysis showed that heart rate (OR 1.054; CI 1.014-1.095), hemoglobin (OR 1.878; CI 1.830-1.928), erythrocyte distribution width (OR 1.171; CI 0.977-1.402), erosion degree of ulcer surface (A and B) (OR 1.191; CI 1.051-1.766), blood transfusion intervention treatment (OR 12.296; CI 1.892-79.917), were a risk factor for early re-bleeding in peptic ulcer bleeding patients in both groups. This study concluded that heart rate was an independent risk factor for early re-bleeding in patients with peptic ulcer and bleeding. Heart rate refers to the number of heartbeat per minute in normal people’s quiet state, which is generally 60 ∼ 100 times/min. It varies from person to person depending on age, sex, or other biological factors. Changes in heart rate and blood pressure are important indicators of blood loss. In acute gastrointestinal bleeding, the blood volume decreases, and the heart rate increases when the body compensates. If the blood volume cannot be stopped or replenished in time, the pulse is weak in a shock state. Heart rate is an indicator of early compensation and early disease progression in patients with upper digestive tract bleeding. This study also shows that hemoglobin is an independent risk factor for early re-bleeding in patients with peptic ulcer and hemorrhage. A domestic report showed that low hemoglobin levels are associated with the prognosis of acute non-variceal upper gastrointestinal bleeding (ANVUGIB) [7]. Other studies have shown that hemoglobin <8g/ dL is a risk factor for re-bleeding in peptic ulcer bleeding (PUB) patients [20]. Some studies conducted multivariate analysis on 897 PUB patients, and one of the risk factors for re-bleeding was reduced hemoglobin [21]. Xu Yongju et al. also reported that hemoglobin content and endoscopic Forrest classification were the main influencing factors for re-bleeding in patients [7]. There have also been related reports abroad confirming that hemoglobin is one of the risk factors for peptic ulcer re-bleeding [22–23]. The reason for this is that the low hemoglobin level in PUB patients indicates a large amount of blood loss and also indirectly indicates the involvement of blood vessels in ulcers. However, when digestive tract bleeding occurs in PUB patients, hemoglobin cannot accurately reflect the severity of bleeding, and factors such as blood concentration should be considered. RDW indicates the degree of red blood cell volume heterogeneity and has been used to identify the type of anemia. Recent studies have shown a certain correlation between RDW and gastrointestinal bleeding a study on the characteristics and etiology of blood cell examination in 170 patients with no result of RDW found that increased RDW was associated with gastrointestinal bleeding [24]. Ryong Lee, [25] revealed that higher RDW levels (especially more than 14.5%) were closely associated with high-risk UGIB patients. There are few reports in relevant literature at home and abroad. This study showed that RDW was an independent risk factor for early re-bleeding in patients with peptic ulcer and hemorrhage. Existing studies have shown that there may be the following reasons for the increase in RDW: Firstly, the patients with PUB might be treated with fasting after admission on account of intermittent hematemesis and hematochezia. Hence, nutritional deficiency was one of the reasons for the increase in RDW [26]. Secondly, Erythropoietin (EPO) is the main factor that causes the increase of RDW. Massive bleeding in the gastrointestinal tract might trigger the physical blood volume to decline, which may stimulate the physical body to release EPO and stimulate the production of red blood cells and HGB. Simultaneously, high levels of EPO can lead to a cascade of reactions. Moreover in the process of bleeding, due to the decrease of blood volume, hemoglobin will drop sharply, stimulating the bone marrow to produce a large number of hetero-erythrocytes [27]. Finally, other studies have shown that vascular inflammatory reaction [28], severe mental stress [29], oxidative stress, and imbalance of intestinal microflora can all boost the RDW value in patients with bleeding [30]. With the booming economy and rising living standards, the incidence of chronic ailments increases year by year, especially for cardiovascular and cerebrovascular diseases develops. Consequently, patients with PUB resulting from NSAID intake on account of cardiovascular ailments have increased. Preliminary studies revealed mucosal lesions in 30%-50% of patients with NSAIDs [31].The endoscopic features of NSAIDS-related ulcers are as follows: most of the ulcers are found in the gastric antrum, but also in other parts of the stomach. The appearance and size of the ulcers are different, mainly multiple shallow ulcers [32, 33]. Some patients with Forrest III peptic ulcer have a clean base of ulcer but there is erosion around the ulcer and a tendency to bleed, or even to bleed heavily. This study showed that the extent of ulcer erosion is an independent risk factor for early re-bleeding in patients with peptic ulcer bleeding. The mechanism of bleeding induced by drug-related ulcers may be as follows: Firstly NSAIDs are weakly acidic lipid-soluble drugs, which enter cells through the mucosal system, and acidify cells; it can facilitate the permeability of epithelial mucosal cells and the diffusion and transport of hydrogen ions, destroying the stability of bicarbonate barrier. In addition, after NSAIDs enter the blood circulation, they can inhibit the activity of cyclooxygenase-1, reduce the synthesis of prostaglandin, suppress the blood supply of gastric mucosa, and affect the repair and reconstruction of gastric mucosa, leading to mucosal disorder and the formation of ulcers [34]. Because, the Forrest grade, CRS score and GBS score did not involve mucosal surface degree around ulcer. In this study, it was concluded that the degree of erosion ulcer surface: A indicating red surface, B indicating black surface were independent risk factors for early re-bleeding of peptic ulcer. Therefore, it is necessary to give more attention to the characteristic of the erosion surface the ulcer during clinical endoscopy, early judgment of the patients re-bleeding and degree of risk with provide reference for clinical treatment and improve the patients prognosis. If the blood volume of patients with peptic ulcer bleeding is greater than 400ml, Symptoms such as: dizziness, palpitation, fatigue and etc may occur. If blood loss is greater than 1000ml within short period of time shock may appear. This study demonstrates that transfusion intervention is an independent risk factor for early re-bleeding in patients with peptic ulcer and bleeding. Different studies have reported that about 35-40% of UGIB patients received at least one transfusion of red blood cell suspension to correct anemia during intervention treatment [35]. Overall, 149/642 (23.2%) patients received transfusion. In adjusted models, transfusion was associated with higher odds of early re-bleeding (report OR and 95% CI here).

### Clinical implications

The guidelines and textbooks point out that the indications for infusion of concentrated red blood cells are: SBP is less than 90mmHg, or a reduction of more than 30mmHg from the baseline SBP; Increased heart rate (≥ 120beats/min), Hemoglobin less than 70g/L or RBC less than 25%. However, consistent with current guidelines, when patients have clear heart disease or hemodynamic instability, it is recommended that the blood transfusion indications be relaxed than before. In the previous study, at the same threshold, the number of patients with cardiovascular disease who received blood transfusion by clinicians was twice as high as that without intervention (50%) [35]. A meta-analysis included only five randomized controlled trials in patients with acute gastrointestinal bleeding, showing that restrictive blood transfusion strategies are associated with lower all-cause mortality and re-bleeding [36]. NICE guidelines recommend higher blood transfusion levels in patients with cardiovascular disease than in patients without cardiovascular disease, and recommend a hemoglobin threshold of 80g/L in patients with cardiovascular disease and 70g/L in patients without cardiovascular disease [37]. The etiology of re-bleeding in PUB patients is complex, and individualized blood transfusion management methods are needed. Timely, guideline-concordant transfusion strategies may reduce adverse events in PUB; however, practice should follow restrictive thresholds where appropriate [38]. The Forrest classification – used for >40 years—stratifies endoscopic stigmata of recent hemorrhage into categories that inform re-bleeding risk and choice of hemostasis. Its clinical role has been repeatedly validated [38, 39]. Forrest classification divides the signs of bleeding under gastroscopy into different levels which is helps to judge of re-bleeding, so as to make a more objective evaluation of the choice of endoscopic treatment methods and the efficacy of clinical treatment methods. Studies shows [13] that in patients with peptic ulcer bleeding, Forrest grading is associated with the recurring of bleeding, applicable to clinical and as the basis of choosing endoscopic hemostatic method over gastroscopy, but different mortality rates compare Forrest classifying patients that there was no statistical difference, then corresponding an increase in bleeding rate and no alteration in the mortality rate. Other studies show that Forrest grade has a predictive value for peptic ulcer re-bleeding, which is significant in predicting gastric and duodenal ulcers re-bleeding but does not have the ability to determine its mortality rate [38]. In our study the risk of re-bleeding is higher in Forrest IB 20% compared to other grades (IA 16.66%, IIA8.57%, IIB3.31%, IIC1.92%) and is tendency to re-bleeding again. Oozing hemorrhagic clots are usually formed following massive bleeding due to erosion of large vessels on the ulcer base. These clots temporarily halt the bleeding but with time, they get washed away leading to recurrence of bleeding. Linda zhang [6] is consistent with the results Forest grading IIb is associated with peptic ulcer bleeding and bleeding in patients with hemorrhage. It is advisable that UGIB patients visit clinics with endoscopy facilities for the assessment using Forrest classification to foreshadow re-bleeding risk and undertake preventive measures to prevent major fatal bleeding.

### Limitations

This single-center retrospective study is subject to selection bias and residual confounding (e.g., medications, comorbidities, diet). Some variables had missing data despite complete-case analysis; inter-observer variability in endoscopic grading may persist. We did not perform external validation, and subgroup analyses had limited power.

### Future research

Multicenter, prospective validation with pre-registered analysis and decision-curve evaluation is needed to confirm generalizability and clinical utility [39].

This study retrospectively analyzed the clinical value of Forrest grading, CRS and GBS predicting the risk of early re-bleeding in PUB patients. The results showed that in PUB patients, the Forrest grading and GBS had better predictive ability than the CRS in predicting early post-treatment re-bleeding. Prior works has evaluated how endoscopic stigmata (Forrest) and clinical scores (GBS/CRS) relate to early re-bleeding risk [39, 40]. The aim is to observe the prognostic value of Forrest grade and Rockall score in patients with peptic ulcer hemorrhage. The results show that Forrest grade is correlated with the incidence of re-bleeding in patients with peptic ulcer hemorrhage, while Rockall score is correlated with mortality [13]. This is similar to our research results. Expert consensus (2015) recommends GBS for initial assessment because it relies on clinical and laboratory data and does not require endoscopy; its value for re-bleeding prediction is more limited than Forrest in our cohort [41]. And the sensitivity is high without endoscopy and the judgment for low-risk patients can safely in the outpatient treatment effectively improve the diagnosis and treatment efficiency [42]. However, Zhiyu Dong et al. found no difference between CRS score and GBS score in predicting the risk of re-bleeding in patients with peptic ulcer [43]. Wang Kai et al. [44] believe that Rockall scoring system has a low predictive value for re-bleeding in patients with ANVUGIB, but its predictive ability for re-bleeding in patients with ANVUGIB at different ages is inconsistent. In China, Ran Yan et al. found that Blatchford grading had a high predictive value for re-bleeding in non-elderly patients, but a low predictive value for re-bleeding in elderly patients [45]. In summary, the Forrest grading and GBS scoring system include endoscopic performance of patients, and the data are not easy to obtain. However, most studies have shown that the two scoring systems above have high predictive value for early re-bleeding risk in PBU patients [46]. Blatchford scoring mainly included a number of clinical and laboratory indicators of patients, and its predictive value for early re-bleeding in PBU patients were limited. However, data of this scoring system was easy to obtain and could be used to predict the risk degree of early disease in PUB patients [47].

## CONCLUSION

Heart rate, hemoglobin levels, erythrocyte distribution width, degree of ulcer erosion, and the need for blood transfusion intervention were identified as independent risk factors for early re-bleeding in patients with peptic ulcer bleeding (PUB). Among the scoring systems evaluated, the Forrest grading system demonstrated superior predictive power for early re-bleeding following treatment when compared to the CRS and GBS systems. However, the GBS system also proved valuable for stratifying the risk of re-bleeding in clinical practice. Its utility could be further enhanced by incorporating additional factors such as endoscopy findings and patient age into the scoring criteria.

## AUTHORS’ CONTRIBUTION

Conceived and designed the study; collected the data; checked the follow-up; wrote different parts of the manuscript; revised the manuscript. All authors approved the final manuscript.

## ETHICAL APPROVAL

This study involved a retrospective analysis of fully de-identified medical records. According to the official determination issued by the Ethics Committee of Ningxia Medical University, the project “uses fully de-identified data extracted from existing medical records, involves no direct or indirect patient contact, no intervention, and no re-identification, and poses no more than minimal risk.” The Committee therefore determined that the study is EXEMPT from full ethics review under the University’s criteria for retrospective chart-review research using de-identified data (Exemption No.: EMNU-1124/2025, dated 02 November 2024).

## PATIENT CONSENT

Informed consent was not required. Investigators adhered to all data-security and confidentiality safeguards.

## AVAILABILITY OF DATA AND MATERIAL

The fully de-identified minimal dataset and all analysis code used to produce the results are publicly available at Zenodo (DOI: 10.5281/zenodo.17593134). The deposit includes a data dictionary (variable definitions) and scripts sufficient to reproduce every table and figure without access to the original records. Dataset license: CC0 1.0; code license: MIT. No restrictions apply to data access

## Data Availability

The fully de-identified minimal dataset and all analysis code used to produce the results are publicly available at Zenodo under an open license, DOI: https://doi.org/10.5281/zenodo.17593134. The deposit includes variable definitions and scripts sufficient to reproduce every table and figure without access to the original records.

## ACKNOWLEDGEMENTS

I am grateful to those units and individuals who have given all kinds of support, guidance, and assistance to complete the research work and provided all sorts of favorable conditions for the paperwork.

## GENERATIVE AI DISCLOSURE

Language editing assistance was provided using Grammarly (version **6.8.263**) and ChatGPT (OpenAI; GPT-5 Thinking, accessed [November 2025]), limited strictly to grammar and clarity. No text, analyses, figures, or references were generated de novo. All authors reviewed and take full responsibility for the content.

